# COVID-19 Pandemic Response in a Migrant Farmworker Community: Excess Mortality, Testing Access and Contact Tracing in Immokalee, Florida

**DOI:** 10.1101/2021.10.01.21264382

**Authors:** Neha Limaye, Brennan Ninesling, Frantzso Marcelin, Cody Nolan, Walter Sobba, Matthew Hing, Emily Ptaszek, Fernet Léandre, Daniel Palazuelos

## Abstract

**Introduction:** We aim to estimate the impact of COVID-19 in Immokalee, FL and assess community experiences with workplace conditions, access to testing, sources of information, and contact tracing to inform and strengthen local public health sector efforts in reaching and providing high-quality care to the community.

**Methods:** We conducted a descriptive analysis of data on COVID-19 deaths for Collier County from May-August 2020. We surveyed a cross-sectional, randomized representative sample of 318 adults living in Immokalee from March-November 2020 to assess socio-demographics, sources of information, ability to follow guidelines, and experiences with local programs. Results were compared across language groups.

**Results:** Average excess mortality in Collier County was 108%. The majority surveyed in Immokalee had socio-demographic factors associated with higher COVID risk. Non-English speakers had higher workplace risk due to less ability to work from home. Haitian Creole speakers were less likely to be tested, though all participants were willing to get symptomatic testing and quarantine. Those participants who tested positive or had COVID-19 exposures had low engagement with the contact tracing program, and Spanish-speakers reported lower quality of contact tracing than English speakers.

**Conclusions:** The community of Immokalee, FL is a vulnerable population that suffered disproportionate deaths from COVID-19. This study reveals language inequities in COVID testing and contact tracing should be targeted in future pandemic response in Immokalee and other migrant farmworker communities.

## 1. Introduction

### 1.1. Background

Migrant and seasonal farmworkers (MSFW) face enormous structural barriers to health, including poverty, food insecurity, poor working conditions, high occupational hazard, and limited access to healthcare.^1^ Although data are lacking, those available suggest that MSFW experience inequities in multiple health outcomes.^2–4^ At work they face challenging environmental conditions including heat, sun, pesticide exposure, and dust and particle inhalation.^1^ Additionally, most MSFW are uninsured and must overcome language barriers, long work hours, and lack of transportation to attend health appointments, with care often only available at Federally Qualified Health Centers (FQHC).^5^ Consequently, the majority of MSFW do not have primary care physicians.^6^

The COVID-19 pandemic has only deepened pre-existing health inequities. Data demonstrate disproportionate rates of COVID-19 morbidity and mortality among Black, Latinx and immigrant populations, who make up the majority of MSFW in the U.S.^7–9^ MSFW are at especially high risk for COVID-19 exposure and subsequent infection due to high density housing and ‘essential worker’ status, which has exempted them from the protection of working from home enjoyed by other industries.^10,11^ Typical working conditions are not conducive to distancing, as many MSFW work side-by-side and share transportation to and from work.^12^ Once infected, spread within MSFW communities is facilitated by lack of testing and contact tracing, and by housing conditions which preclude distancing or effective quarantine.^13^ Finally, those infected are at higher risk of severe COVID-19 disease due to underlying health conditions that predispose to morbidity and mortality.

Although the plight of MSFW during the COVID-19 pandemic has gained media attention, there is minimal data around its true impact. Access to COVID-19 testing in this population is unknown, and the true burden of death due to COVID-19 in these communities remains unclear. Contact tracing is a key aspect of controlling COVID-19 spread and is especially important for MSFW, who move around frequently for work. However, no studies have examined the success or quality of contact tracing programs in an MSFW population. Data around COVID-19’s specific impact on MSFW communities in Florida are lacking, despite Florida housing one of the largest populations of MSFW in the country.

### 1.2. Study Site: Immokalee, Florida

Immokalee is a rural community located in southwest Florida’s Collier County, the center of the nation’s tomato growing industry. Much of the population, from 20,000 to 27,000 residents, works in agriculture as MSFWs or packing house workers, with 37.4% living below the poverty line. Residents are primarily from Mexico, Central America, and Haiti; 68% of the population speak Spanish at home and 13% speak Haitian Creole.^6^ Occupational, economic, and linguistic factors confer substantial vulnerability to COVID-19 infection in Immokalee; despite its small size, Immokalee was the Florida zip code with the highest number of COVID-19 cases in the state in June 2020.^14^ Positivity rates at the time were as high as 36% compared to 5.6% in the state.^15^ After observing disease burden and difficulties in accessing COVID-19 testing and contract tracing among MSFWs, the Coalition of Immokalee Workers (CIW) — a local human rights organization —facilitated partnerships between the Collier County Department of Health (DOH), Partners in Health (PIH), Doctors Without Borders (Medicins Sans Frontieres - MSF), and a local FQHC (Collier Health Services, Inc., d/b/a Healthcare Network (HCN)). The DOH led all contract tracing efforts in Immokalee, and offered nasal PCR testing by appointment until November 2020, with results given via an English-language online portal which asked for a social security number. Starting in November 2020, HCN began offering walk-up rapid testing in Immokalee, with results given within an hour by Spanish- and Haitian Creole-speaking staff. Both HCN and DOH created community health worker programs for COVID-19 outreach, but all positive results from HCN’s testing were forwarded to the DOH’s contact tracing system (see Appendix A, Figure 1 for timeline).

In this study, we first aimed to estimate the impact of COVID-19 on Immokalee, FL by calculating excess mortality from publicly available data. Then, we conducted a household survey to collect sociodemographic information and assess community experiences with workplace conditions, access to testing, sources of information, and the DOH contact tracing program.

## 2. Methods

### 2.1. Descriptive Analysis

We collated data from the Florida Department of Health and Medical Examiners Offices for Collier, Lee, Hendry, Glades and Orange Counties. First, we tabulated the total deaths from COVID-19 for residents of Collier County. Data for the Immokalee zip code alone were not available. We then compared deaths for Collier County during May-August 2015-2019 with deaths from May-August 2020. Excess mortality was calculated in accordance with CDC guidelines, where death data were grouped weekly to account for temporal effects. Excess mortality was calculated for each week and then summed together to find excess mortality for the period beginning 4/27/20 and ending 8/16/20. Data were disaggregated by age (under 60 or ≥60 years) and sex.

### 2.2. Questionnaire

The questionnaire was created using demographic and social screening tools from the National Agricultural Workers Survey and various farmworker healthcare organizations.^7–9^ Spanish- and Haitian Creole-speaking CIW staff reviewed the tool for understandability by the local population. The questionnaire (Appendix B) assessed the following thematic areas:

- Demographic and socioeconomic information
- Sources of news and information on COVID-19
- Ability to follow COVID-19 precautions
- Experiences with contact tracing

### 2.3. Sample Size

Our target population was adults living in Immokalee during the months of March-November 2020. We estimated a population of 12,000 adults in Immokalee, a 95% confidence interval, a precision of 5%, and a prevalence of 15-20% of the population meeting requirements to be contacted by contact tracers (given an estimated local COVID-19 prevalence of 8%). From our initial sample size of 193-241 participants, we accounted for intercorrelation in order to survey multiple people per household. We used an intracluster correlation (ICC) of 0.1-0.33 and an estimated household size (m) of ∼4 to calculate a design effect of 1.3-2, giving us a final sample size goal of 300-350 participants.

We used a publicly available address list from the Collier County Property Appraiser’s office and extracted 350 addresses using a random number generator. Study staff visited each address, and after exhausting that first address list, extracted 100 further addresses at a time to reach a sample size of 300-350 participants.

### 2.4. Study Procedures

From January 18 - March 11, 2021, study staff (FM and BN) visited addresses on weekends and evenings in a predetermined fashion considering geographical location. They wore appropriate personal protective equipment at all times and followed social distancing guidelines. They conducted multiple surveys per household as household members may have had different experiences, and it is common for multiple families in Immokalee to cohabitate.

Study staff started each visit with an initial screening to confirm each participant 1) was 18 years or older, 2) lived in Immokalee, and 3) lived in Immokalee for at least 2 weeks between March-November 2020. If inclusion criteria were met, study staff obtained verbal informed consent in the participants’ preferred language. Verbal consent was obtained due to the population’s literacy level and to maintain participant anonymity. If participants preferred to participate at a different time or no adults were home, one follow-up visit was done at the same address. All households visited received a paper pamphlet detailing available health resources in their preferred language.

Questionnaire responses were recorded in REDCap.^16,17^ The study staff entering data were trained on how to use REDCap, and data were checked weekly by the co-investigators to ensure internal validity. The study was reviewed by the Institutional Review Boards at Mass General Brigham and the Harvard School of Public Health and met criteria for exemption.

### 2.5. Data Analyses

All analyses were conducted utilizing R (Version 4.0.2, The R Foundation, 2021) and Rstudio (Version 1.3.959, RStudio Team, 2021). Chi-square and Fisher’s exact tests were utilized to compare variables as appropriate. Of note, data from a question on sick leave was not analyzed due to participant misinterpretation of the question.

## 3. Results

### 3.1. Descriptive Analysis

An analysis of Collier County mortality data from April 27^th^ through August 16^th^ revealed an average excess mortality of 108% (167 excess deaths). When data were disaggregated by sex alone, age alone, and both sex and age, excess mortality findings were largely consistent (107%, 115%, and 115%, respectively, see Appendix A, Table 1).

### 3.2. Study Population

Of 550 households randomized, 131 did not answer the door, 140 were not interested in participating, and 279 agreed to participate. From these, 318 individuals consented to participate and were surveyed (Appendix A, Figure 2).

Baseline participant and household demographics are depicted in Table 1. Spanish (42.1%) was the most frequent preferred language, with English (37.3%) and Haitian Creole (18.7%) accounting for most other participants. Nearly one-fifth of participants reported food insecurity during the past month (19.2%). The mean household size was 3.95 persons with homes averaging 0.856 bedrooms per person and 0.483 bathrooms per person.

**Table 1.**
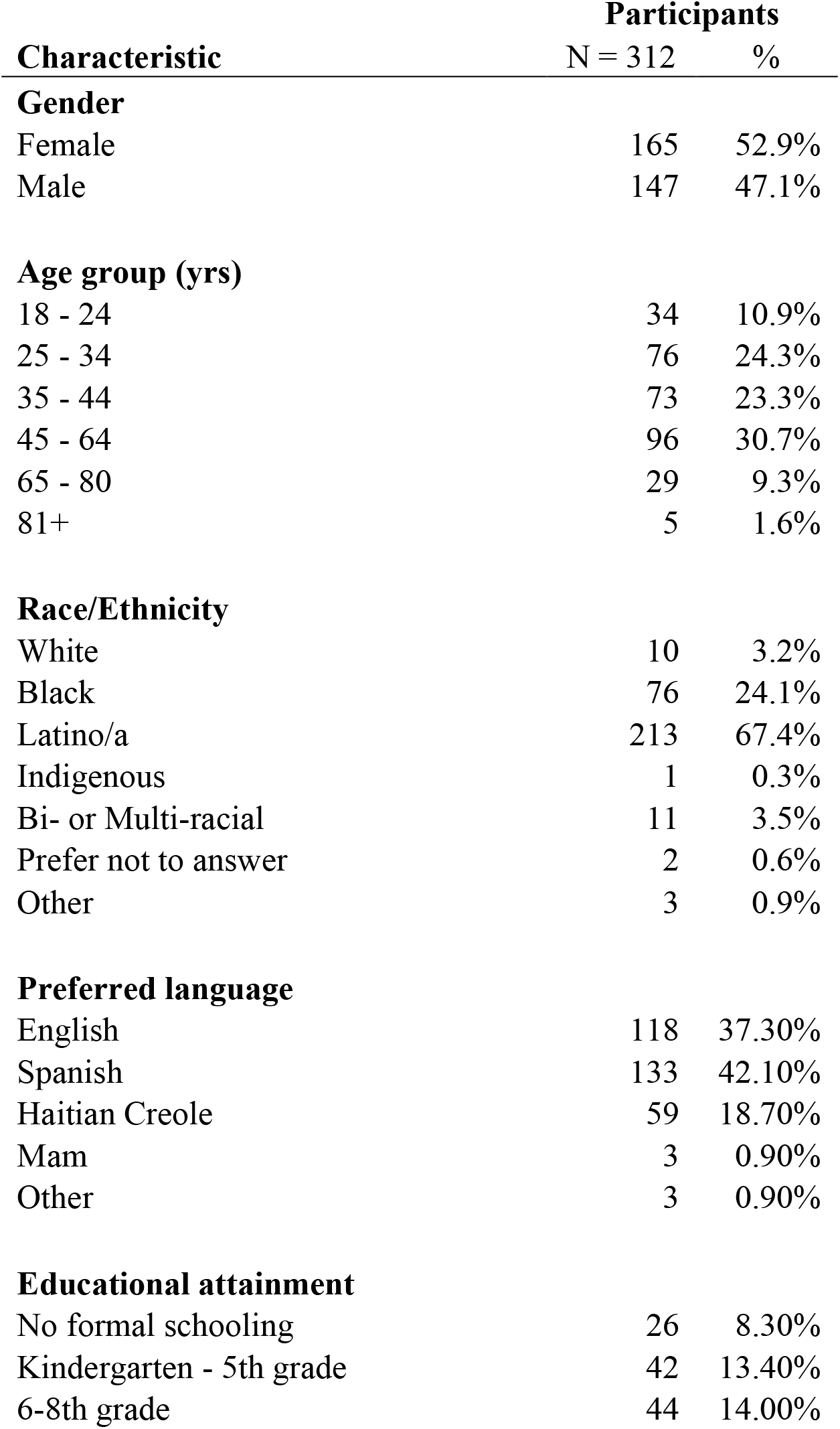
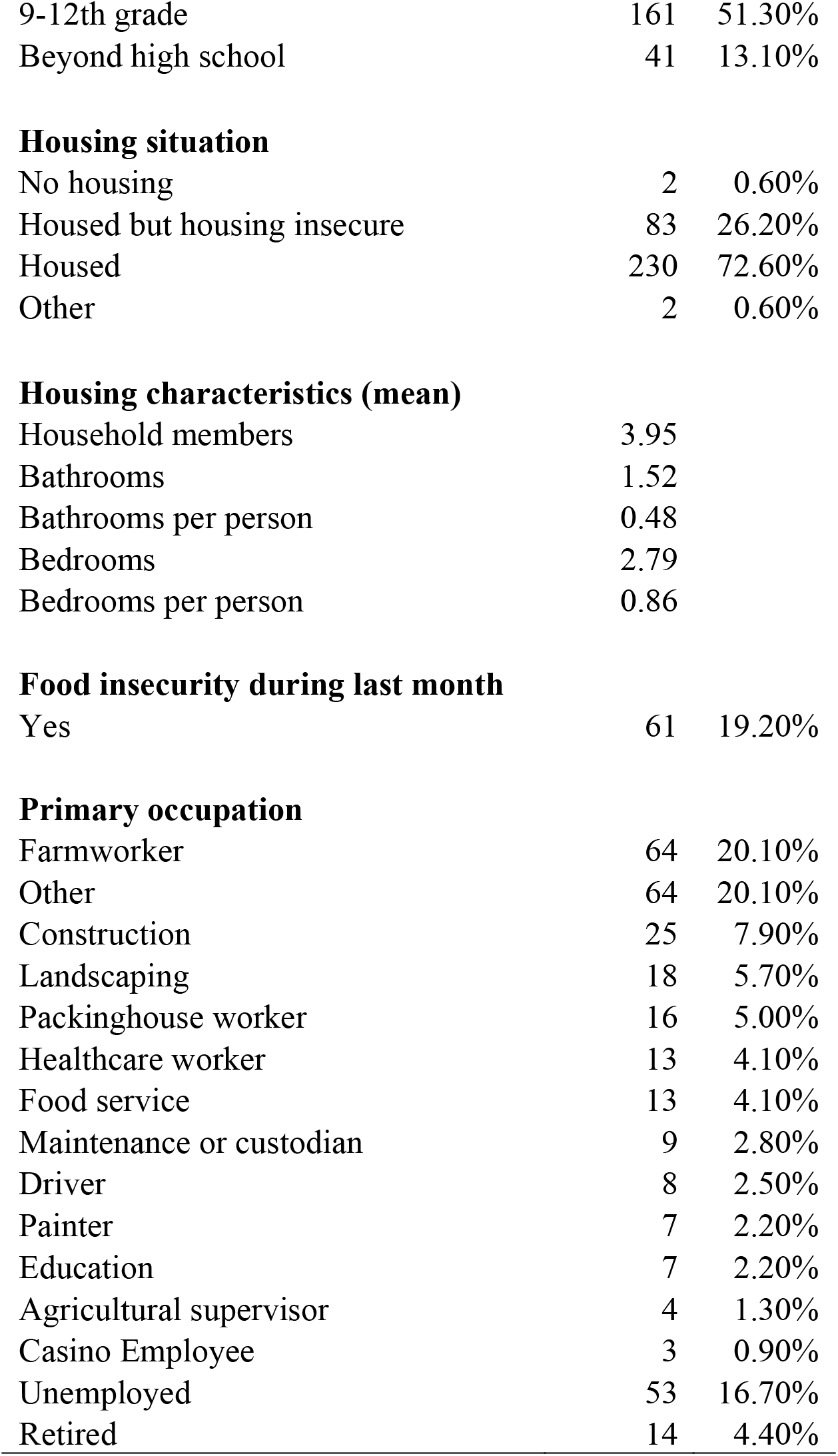
Demographic characteristics of survey participants

| **Characteristic** | **Participants** |  |
| --- | --- | --- |
|  | **N = 312** | **%** |
| **Gender** |  |  |
| Female | 165 | 52.9% |
| Male | 147 | 47.1% |
| **Age group (yrs)** |  |  |
| 18 - 24 | 34 | 10.9% |
| 25 - 34 | 76 | 24.3% |
| 35 - 44 | 73 | 23.3% |
| 45 - 64 | 96 | 30.7% |
| 65 - 80 | 29 | 9.3% |
| 81+ | 5 | 1.6% |
| **Race/Ethnicity** |  |  |
| White | 10 | 3.2% |
| Black | 76 | 24.1% |
| Latino/a | 213 | 67.4% |
| Indigenous | 1 | 0.3% |
| Bi- or Multi-racial | 11 | 3.5% |
| Prefer not to answer | 2 | 0.6% |
| Other | 3 | 0.9% |
| **Preferred language** |  |  |
| English | 118 | 37.30% |
| Spanish | 133 | 42.10% |
| Haitian Creole | 59 | 18.70% |
| Mam | 3 | 0.90% |
| Other | 3 | 0.90% |
| **Educational attainment** |  |  |
| No formal schooling | 26 | 8.30% |
| Kindergarten - 5th grade | 42 | 13.40% |
| 6-8th grade | 44 | 14.00% |
| 9-12th grade | 161 | 51.30% |
| Beyond high school | 41 | 13.10% |
| **Housing situation** |  |  |
| No housing | 2 | 0.60% |
| Housed but housing insecure | 83 | 26.20% |
| Housed | 230 | 72.60% |
| Other | 2 | 0.60% |

**Housing characteristics (mean)**
|  |  |
| --- | --- |
| Household members | 3.95 |
| Bathrooms | 1.52 |
| Bathrooms per person | 0.48 |
| Bedrooms | 2.79 |
| Bedrooms per person | 0.86 |
| **Food insecurity during last month** |  |  |
| Yes | 61 | 19.20% |
| **Primary occupation** |  |  |
| Farmworker | 64 | 20.10% |
| Other | 64 | 20.10% |
| Construction | 25 | 7.90% |
| Landscaping | 18 | 5.70% |
| Packinghouse worker | 16 | 5.00% |
| Healthcare worker | 13 | 4.10% |
| Food service | 13 | 4.10% |
| Maintenance or custodian | 9 | 2.80% |
| Driver | 8 | 2.50% |
| Painter | 7 | 2.20% |
| Education | 7 | 2.20% |
| Agricultural supervisor | 4 | 1.30% |
| Casino Employee | 3 | 0.90% |
| Unemployed | 53 | 16.70% |
| Retired | 14 | 4.40% |

### 3.3. Essential Worker Status and Workplace Policies

Table 2 details participants’ occupational status and policies. Twenty-six percent of English speakers stated they were offered the option to work from home during the pandemic. By contrast, only 3% of both Spanish and Haitian Creole speakers reported being given the option to work from home (p<0.001 English vs Spanish; p=0.0023 English vs Haitian Creole). The majority of employed individuals in all language groups were categorized as essential workers by the state of Florida.^18^

**Table 2.** Self-reported workplace policies of participants and essential worker status, stratified by primary language spoken

| Survey question | Answer | Primary language spoken |  |  |  |  |  | Comparison |  |
| --- | --- | --- | --- | --- | --- | --- | --- | --- | --- |
|  |  | English |  | Spanish |  | Haitian Creole |  | English / Spanish (*p*) | English / Creole (*p*) |
|  |  | % | No. | % | No. | % | No. |  |  |
| Option to work from home? | No | 74.40% | 67 | 97.00% | 96 | 97.10% | 34 |  |  |
|  | Yes | 25.60% | 23 | 3.00% | 3 | 2.90% | 1 | **<0.0000** | **0.0023** |
| PPE at work? | No | 15.90% | 14 | 26.50% | 26 | 19.40% | 7 |  |  |
|  | Yes | 84.10% | 74 | 73.50% | 72 | 80.60% | 29 | 0.0784 | 0.6388 |
| **Essential worker** |  |  |  |  |  |  |  |  |  |
|  | Yes | 68.60% | 81 | 76.70% | 102 | 66.10% | 39 |  |  |
|  | No | 31.40% | 37 | 23.30% | 31 | 33.90% | 20 | 0.1522 | 0.7329 |

Note: Boldface indicates statistical significance ( $p < 0.05$ ).

### 3.4. COVID-19 Testing Experiences

Table 3 shows participants’ testing experiences, compared by preferred language. 38.1%, 48.1%, and 57.6% of English, Spanish, and Haitian Creole speakers, respectively, reported that they had never been tested for COVID-19. Significantly fewer Haitian Creole than English speakers reported being tested (p=0.014). English and Spanish speakers reported being tested at similar sites, primarily by the DOH, while Haitian Creole speakers were more likely to report being tested at the HCN Immokalee Clinic or HCN-affiliated mobile testing sites (p<.0001). English speakers were more likely to report being tested at sites outside of Immokalee, namely Fort Myers and Naples, compared to Spanish and Haitian Creole speakers.

**Table 3.**
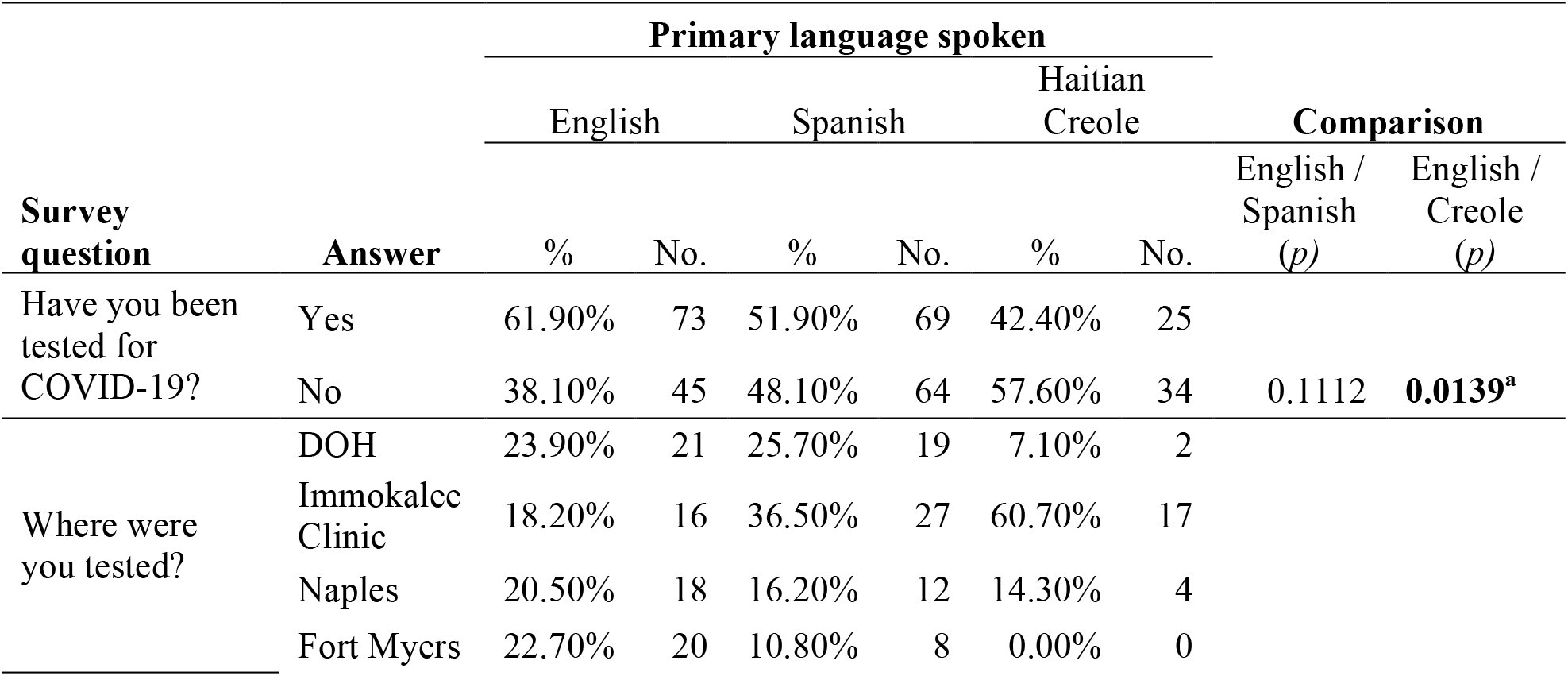
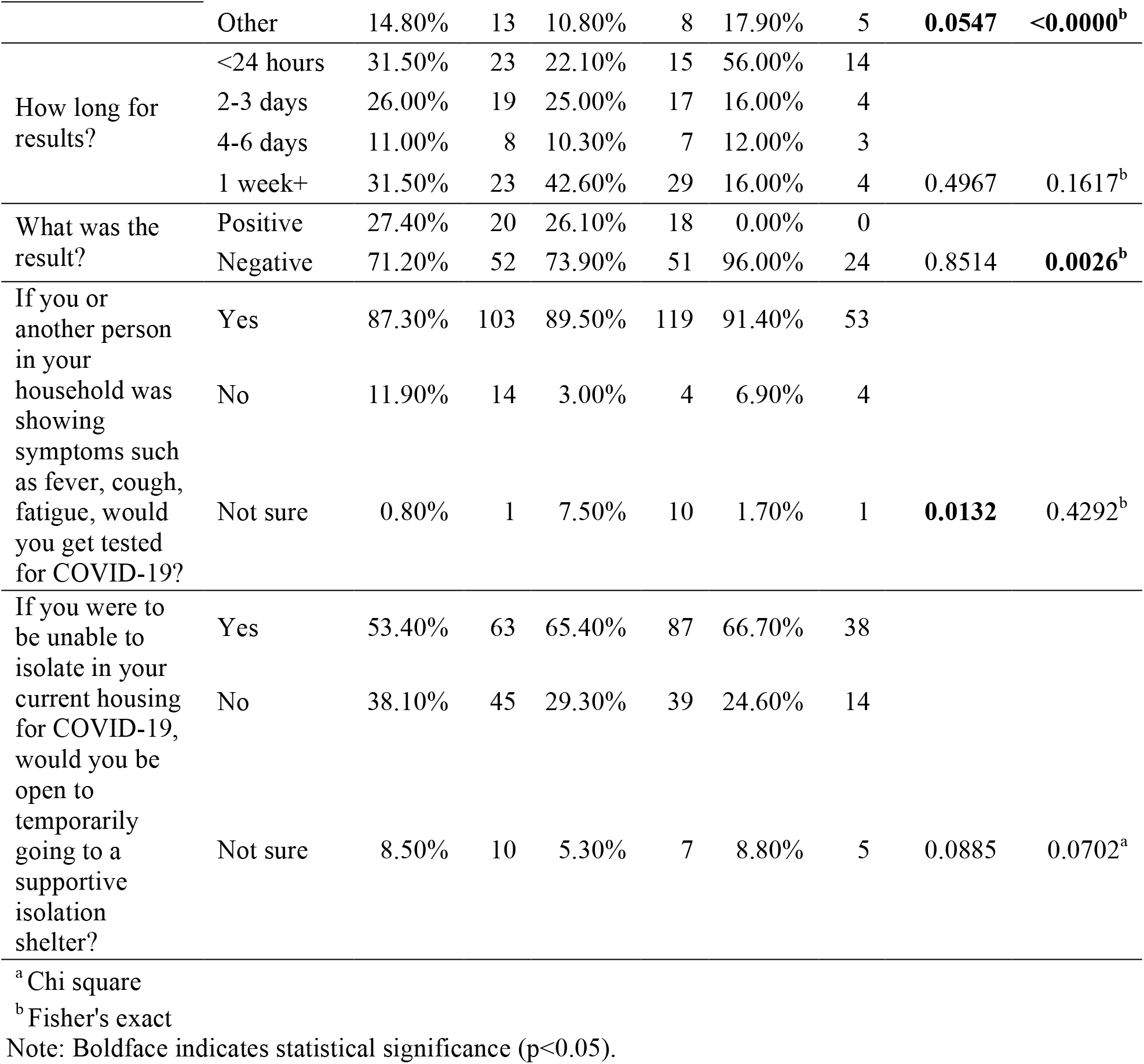
Participant experiences with COVID-19 testing, stratified by primary language spoken

There were no differences in the length of time that lapsed before results were available, though 31.5% of English and 42.6 % of Spanish speakers reported waiting one week or more before receiving their results. More positive COVID-19 test results were reported for English-speaking participants (27.4%) than Haitian Creole-speaking participants (0%), though no significant difference was reported between English and Spanish speaking (26.1%) participants. When asked whether participants would utilize testing resources if exposed to COVID-19, a large share of Spanish (89.5%), Haitian Creole (91.4%) and English speakers (87.3%) reported they would be willing to be tested. The majority of English (53.4%), Spanish (65.4%), and Haitian Creole speakers (66.7%) reported that they would be willing to isolate themselves in a temporary shelter if necessary and they were unable to do so inside their own home.

### 3.5. Quality of Contact Tracing

Those that tested positive for COVID-19 expected to be called by the DOH to trace close contacts, inquire about their ability to quarantine, and provide information about local resources supporting quarantine. Table 4 reflects participants’ experiences with the contact tracing process. No Haitian Creole speakers from our sample reported having tested positive. Only 35% of English speakers and 33% of Spanish speakers who tested positive were asked for names and phone numbers of individuals with which they had been in close contact. 70% of English speakers were asked about their ability to safely quarantine, compared to only 39% of Spanish speakers (p=0.041). Forty-five percent of English speakers reported being provided information on local resources helping with quarantine, compared to 28% of Spanish speakers. Only 26% of English speakers reported being asked about their language preference compared to 82% of Spanish speakers, suggesting that most calls started in English and switched to Spanish if the recipient stated they did not understand English.

**Table 4.**
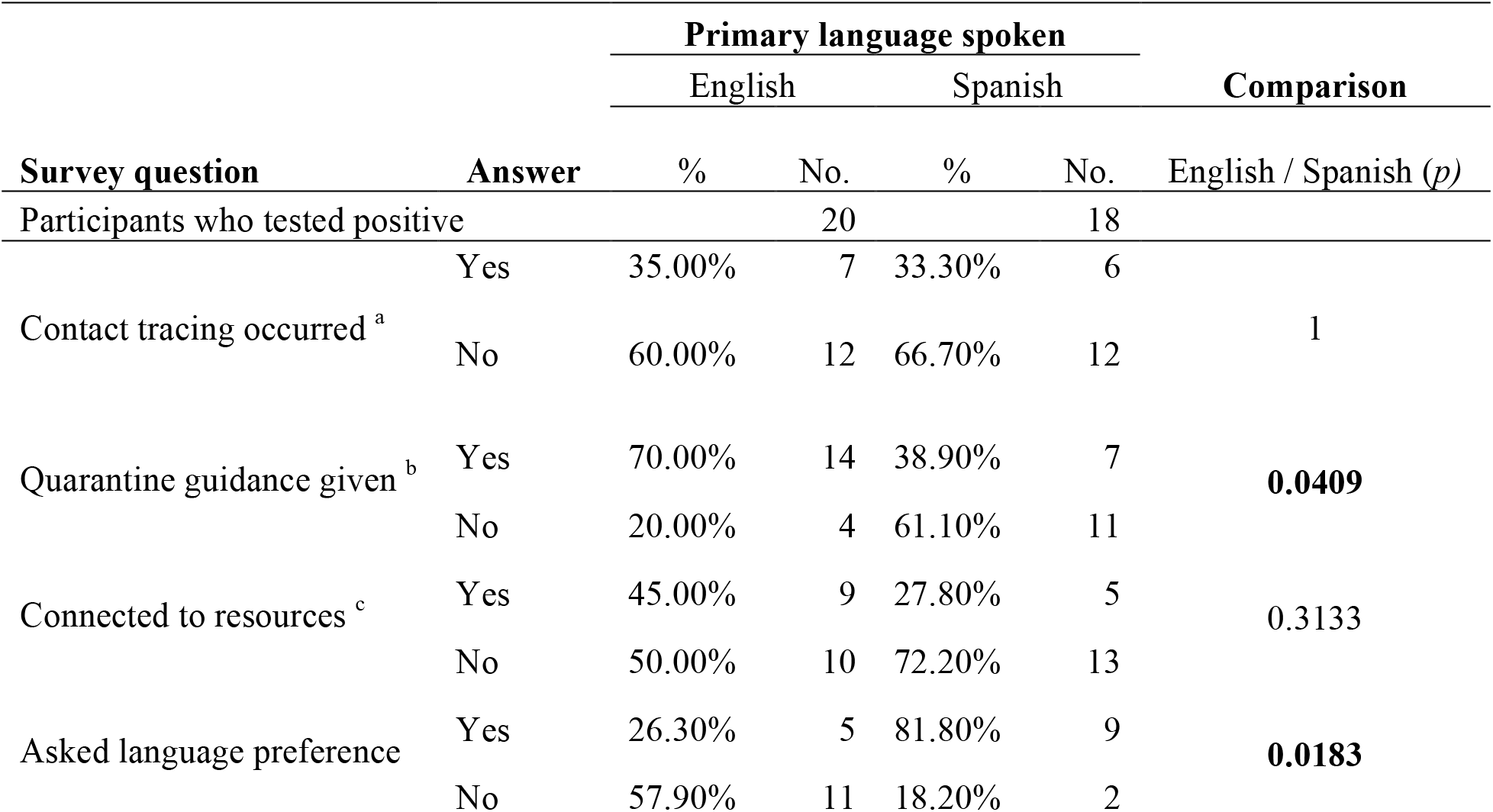
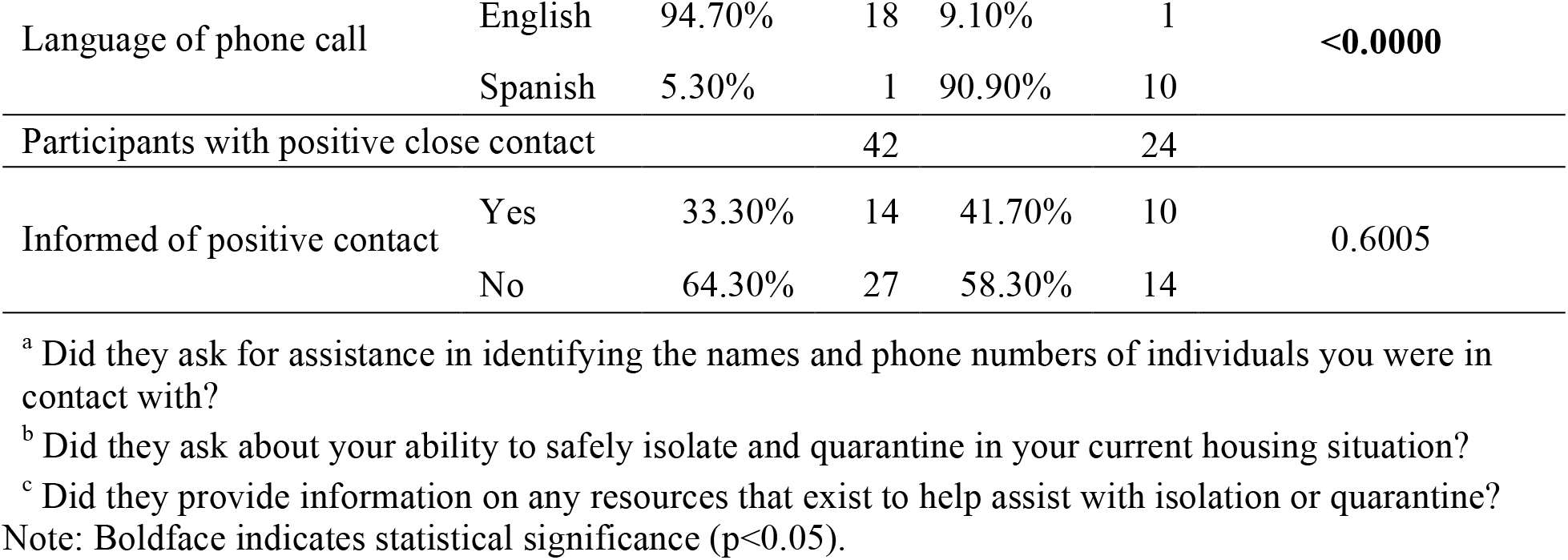
Contact tracing experiences of participants, stratified by primary language spoken

Individuals that were identified as close contacts of someone that had tested positive for COVID-19 were also supposed to be called about their exposure and need to quarantine. 33% of English speakers and 42% of Spanish speakers reported being called by the DOH about their positive close contact.

## 4. Discussion

In this first study of experiences with COVID-19 testing and contact tracing in an MSFW population, we found high excess mortality and high COVID-19 risk, with low testing and contact tracing rates and multiple language-based disparities despite many actions from a coalition of stakeholders. The 108% excess mortality rate in Collier County calculated in this study is extremely high. In comparison, average excess mortality in Florida was estimated to be 15.5% from March to September, with a peak of 38.1% in August,^19^ while nationwide data showed an average of 18.5% excess deaths from March through the end of July.^20^ While we cannot discern how many of these excess deaths were specifically from Immokalee, we know it is an especially vulnerable community within the county, as demonstrated by our data. With an average household size of ∼4 people with shared bathrooms, significant food and housing insecurity, and a preponderance of essential workers, the risk of COVID-19 infection was and continues to be high in this community.^10^ For non-English speakers, that risk is even higher, as our data show they are less frequently able to work from home.

Confronted by this excess disease burden in a highly vulnerable population, the stakeholder groups came together rapidly and worked towards a COVID-19 response. Through their combined efforts, a new system for testing, quarantine and contact tracing was put into place. Our data show the successes and challenges of setting up such a system in Immokalee, with lessons relevant to MSFW populations across the U.S.

The initial testing system was based on appointments scheduled via an English-only online portal. While most surveyed residents indicated willingness to test and quarantine, the data show marked language disparities in testing, with significantly lower testing rates in Haitian Creole speakers. The HCN helped address this disparity a few months into testing by initiating rapid antigen testing at mobile field sites run by Haitian Creole- and Spanish-speaking staff. Our data show that this change may have facilitated a higher testing rate in the Haitian Creole population; amongst the number of Haitian Creole speakers tested, the majority received tests at these HCN rapid-test sites. However, the data show overall low testing rates for all language groups despite these collective efforts, demonstrating a need for even more accessible testing for predominantly MSFW populations.

Our data also highlight challenges with initiating a contact tracing program. Contact tracing is a key strategy for interrupting COVID-19 transmission and providing linkage to quarantine resources for vulnerable individuals,^21,22^ but the percentage of participants in this sample that received contact tracing calls was much lower than the 80% benchmark endorsed by recent studies.^23^ Language disparities were also present in contact tracing: Spanish-speaking individuals were significantly less likely to be asked by contact tracers about their ability to safely isolate.

We were unable to collect any data on Haitian Creole speakers’ experiences with contact tracing, because so few Haitian Creole-speaking respondents were tested. Without access to testing, the true burden of COVID-19 infections in the Haitian population is unknown, and uninterrupted transmission is likely to have occurred.

These disparities in testing and contact tracing highlight the challenges that the population and coalition of stakeholders faced while responding to the high COVID-19 rates in Immokalee. The disparities demonstrate the need for all stakeholders in the larger health system to connect more effectively with vulnerable communities like MSFW, especially Haitian Creole speakers who are known to be particularly marginalized.

To the best of our knowledge, this is the first study to examine community experiences with COVID-19 for MSFW in Florida and the first to evaluate a COVID-19 contact tracing program using probability sampling methods. The population sampled was representative of the diverse Immokalee community with study demographics that parallel 2019 Census Data regarding gender and race distribution and household size.^24^ This study is well-powered to detect differences regarding contact tracing experiences among language groups, especially as we anticipated more clustering than was actually present in our sample.

In terms of future public health responses both in Immokalee and other MSFW communities, it will be key to employ strategies that attend to differential access, vulnerability, and experiences. For example, several programs across the United States indicate promises of linking social support with contact tracing for vulnerable populations and represent an important target for intervention for future health emergencies.^23,25^ The groups working in Immokalee are currently implementing various social support programs, and further research is necessary to understand the effects of this approach. Further research on the Haitian Creole population’s experience will also be essential in planning future responses that better reach the most vulnerable. Moving forward, rapid upscaling of testing access, quality improvement of contact tracing, and community vaccination are needed to prevent continued disproportionate COVID-19 spread and death in vulnerable MSFW populations as new variants of SARS-CoV-2 surge.

### 4.1 Limitations

First, households were sampled from publicly available housing data, which may miss some MSFW living in mobile homes. Also, the survey response rate was just over 50%, raising the possibility of non-response bias. Our suspicion is that non-respondents were more likely to view outreach efforts as intrusive during a pandemic and/or have concerns about migration status; we hypothesize that non-respondents were probably less likely to be reached by public health programs. Additionally, respondents who were migratory from June-October for farm work may have been less likely to receive services. Finally, three participants whose primary language was Mam (a Central American Indigenous language) completed the questionnaire in Spanish, potentially affecting their answers. Future surveys should include all languages in Immokalee.

## 5. Conclusion

Overall, this study quantifies the impact of COVID-19 on Immokalee and elucidates the individual, household, and occupational factors that place this community at especially high risk. The data show that despite coordinated efforts from a committed group of stakeholders, significant language-based inequities impacted the risk of contracting COVID-19, testing rates, and receiving high-quality contact tracing. These inequities are a proxy for the disproportionate barriers faced by non-English speaking populations in Immokalee to access care.

## Data Availability

All data will be available upon request from the authors

## 6. Acknowledgements

The authors would like to thank Marley Monacello and Julia Perkins from the Coalition of Immokalee Workers, John “Trey” Fletcher from the Healthcare Network, and the staff at the Collier County Department of Health for their contributions.

## Author Contributions

**Neha Limaye:** Conceptualization, Methodology, Writing-Original draft preparation, Project administration. **Brennan Ninesling:** Methodology, Investigation, Writing-Original draft preparation, Visualization. **Frantzso Marcelin:** Methodology, Investigation, Writing-Original draft preparation. **Cody Nolan:** Conceptualization, Methodology, Data Curation, Writing-Original draft preparation. **Walter Sobba:** Software, Formal analysis, Writing-Original draft preparation. **Matthew Hing:** Conceptualization, Methodology, Writing-Original draft preparation. **Emily Ptaszek:** Writing-Review & Editing. **Fernet Léandre:** Conceptualization, Writing-Reviewing & Editing. **Dan Palazuelos:** Conceptualization, Methodology, Writing-Review & Editing, Supervision.

The authors declare no conflicts of interest. The authors did not receive funds, grants, or other support from any organization for the submitted work.

## 8. Appendix

**Appendix A, Figure 1.**
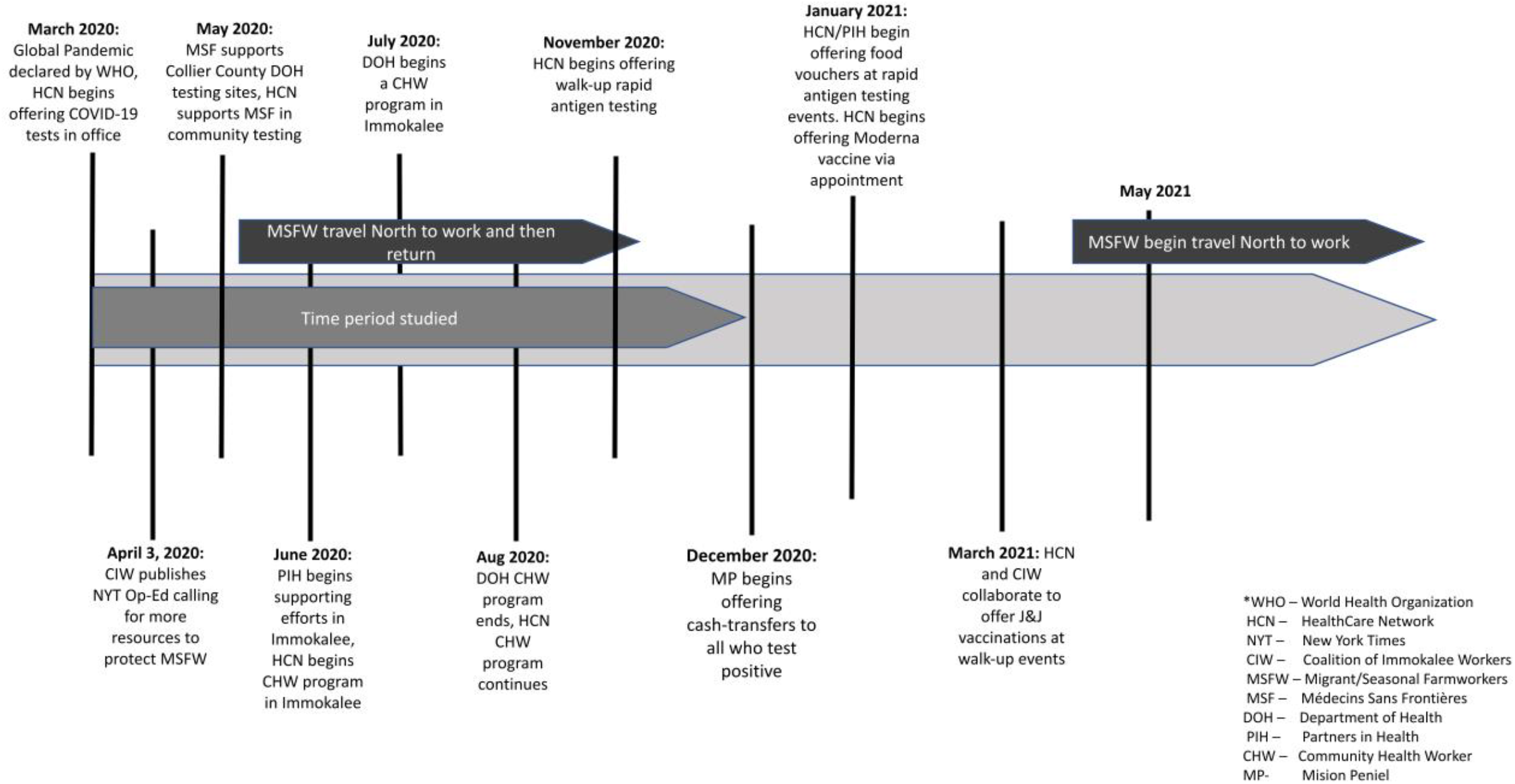
Timeline of COVID-19-related services in Collier County, Florida.

**Appendix A, Figure 2.**
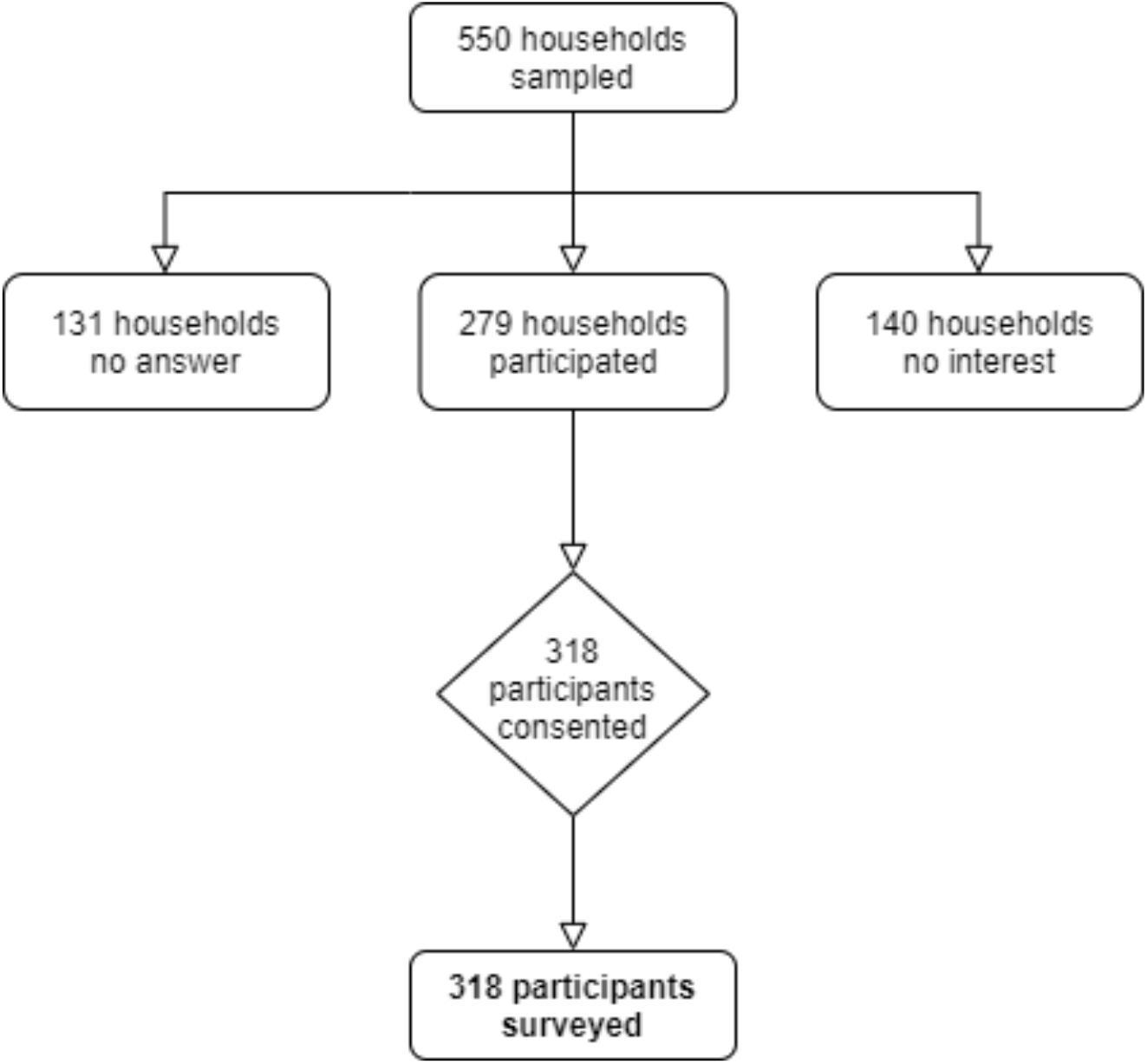
Flow diagram of participant recruitment

**Appendix A, Table 1.**
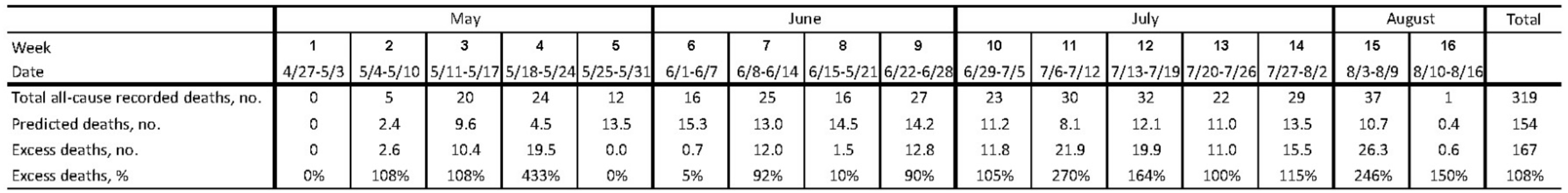
Results for total recorded deaths, predicted deaths, and estimated excess deaths.

## Appendix B

Community Questionnaire, English

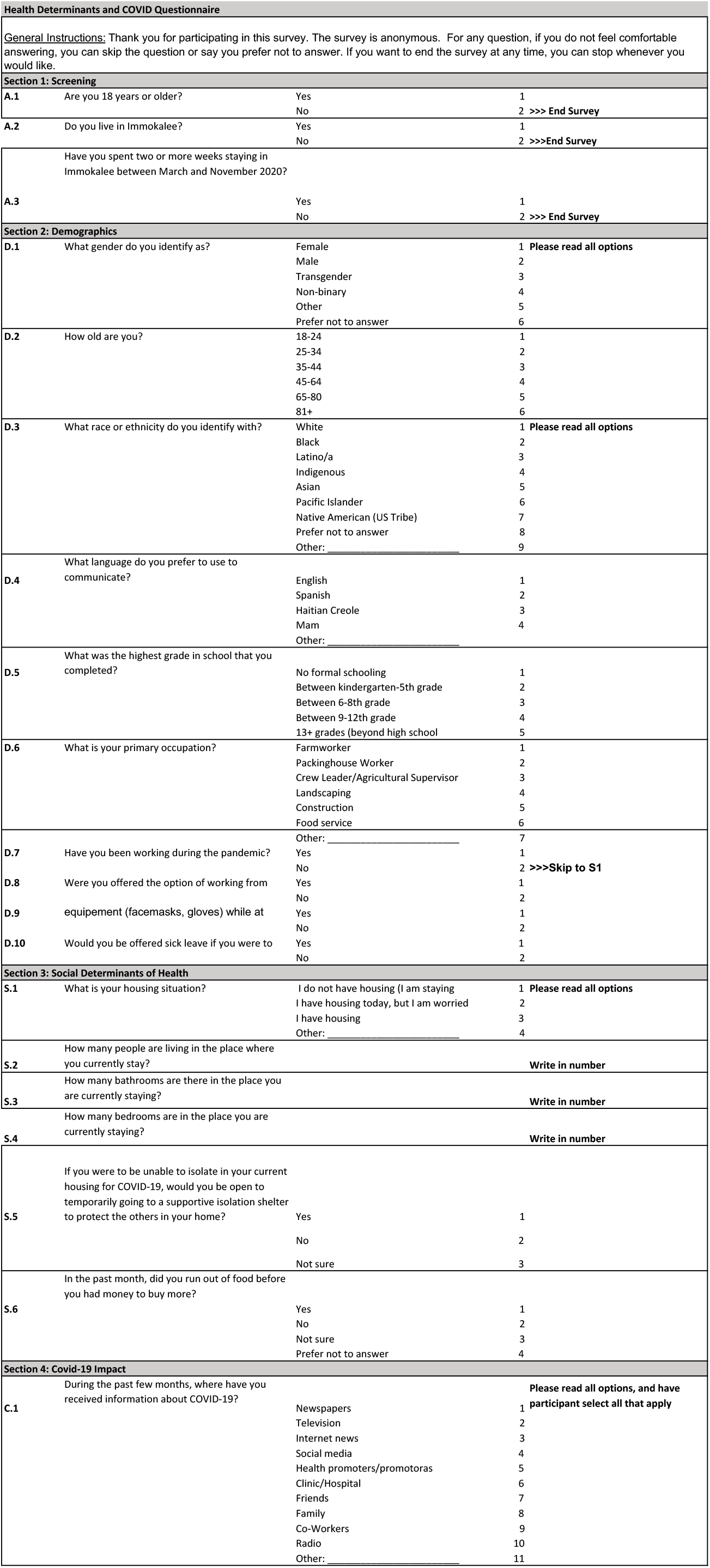
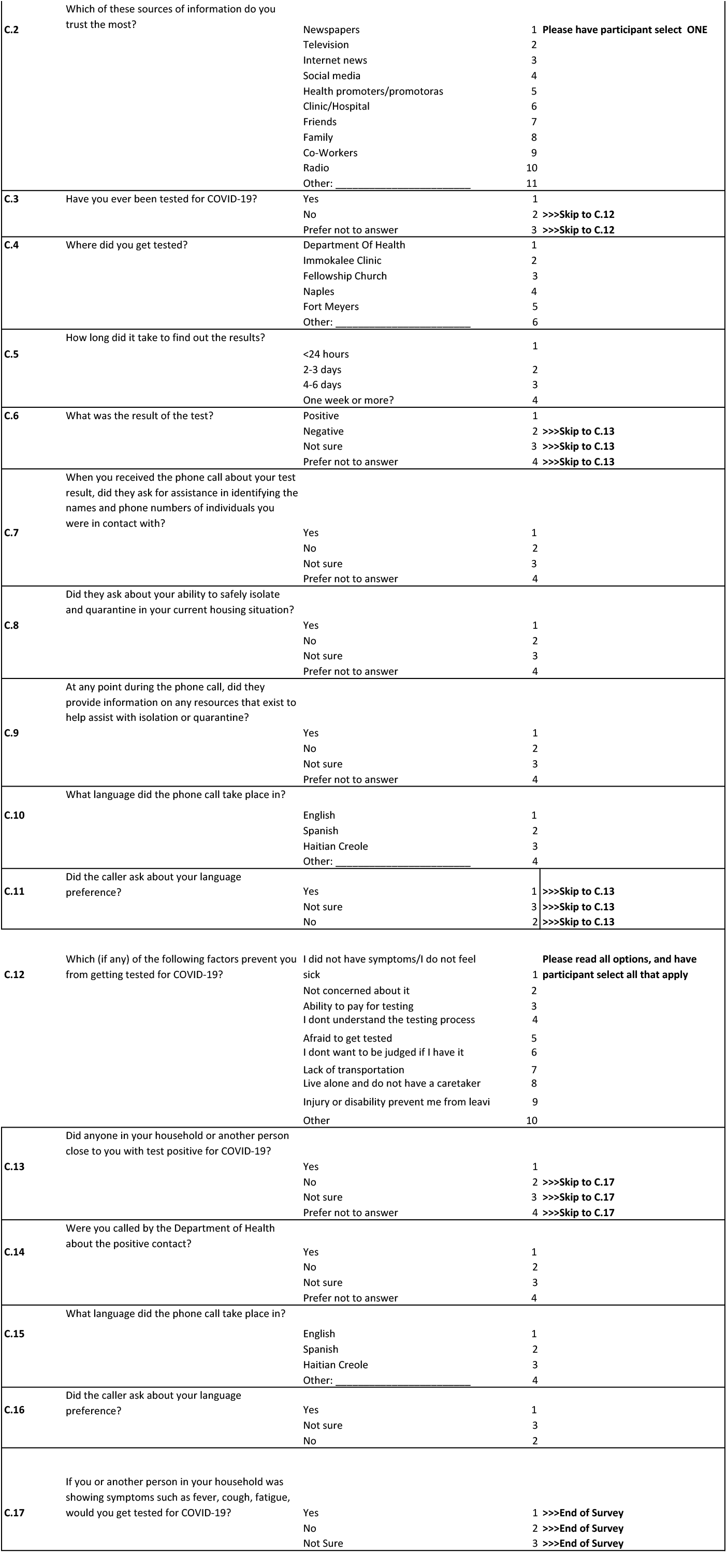

